# Combined Metabolic Activators Improve Cognitive Functions without Altering Motor Scores in Parkinson’s Disease

**DOI:** 10.1101/2021.07.28.21261293

**Authors:** Burak Yulug, Ozlem Altay, Xiangyu Li, Lutfu Hanoglu, Seyda Cankaya, Simon Lam, Hong Yang, Ebru Coskun, Ezgi İdil, Rahim Nogaylar, Ahmet Hacımuftuoglu, Muhammad Arif, Saeed Shoaie, Cheng Zhang, Jens Nielsen, Hasan Turkez, Jan Borén, Mathias Uhlén, Adil Mardinoglu

## Abstract

The neuropathologic hallmarks of Parkinson’s disease (PD) are associated with mitochondrial dysfunction and metabolic abnormalities. We have reported that the Combined Metabolic Activators (CMA), consisting of L-serine, nicotinamide riboside, N-acetyl-L-cysteine, and L-carnitine tartrate can be used in treating metabolic abnormalities. These metabolic activators are the precursors of nicotinamide adenine dinucleotide (NAD^+^) and glutathione (GSH) and used in activation of mitochondrial and global metabolism. We have performed a placebo-controlled, phase-2 study in Alzheimer’s disease (AD) patients and reported that the cognitive functions in AD patients is significantly improved 29% in the CMA group whereas it is improved only 14% in the placebo group after 84 days of CMA administration. Here, we designed a randomized, double-blinded, placebo-controlled, phase-2 study in PD patients with CMA administration. We found that the cognitive functions in PD patients is significantly improved 21% in the CMA group, whereas it is improved only 11% in the placebo group after 84 days of CMA administration. We also found that the administration of CMA did not affect motor functions in PD patients. We performed a comprehensive multi-omics analysis of plasma proteins and metabolites, and revealed the molecular mechanism associated with the treatment of the patients. In conclusion, our results show that treating PD patients with CMAs leads to enhanced cognitive function, as recently reported in AD patients.

## INTRODUCTION

Parkinson’s disease (PD) is characterized by selective degeneration of dopaminergic neurons in the substantia nigra and the presence of fibrillar aggregates, which manifest in motor and non-motor features^1^. The prevalence of PD has surpassed that of Alzheimer’s disease (AD) and many other neurodegenerative diseases^2,3^. Although most PD studies continue to focus on motor endpoints, PD is also being recognized for its complex range of non-motor symptoms^4^, including cognitive impairment, which exist even in the prodromal stages of the disease. Indeed, growing data indicate that metabolic disorders associated with bioenergy failure of nerve cells might increase the risk of developing PD and lead to a higher degree of cognitive impairment and dementia^5^. Moreover, there is considerable evidence for the association between impaired glucose metabolism and PD^6–8^, consistent with a predilection to cortical anaerobic glycolysis in patients with PD^7,9^. Accumulating evidence shows clinical benefits of metabolic treatments in patients with PD (e.g. reduced risk of PD in patients with diabetes using antidiabetics)^10,11^, including some improvement in cognitive decline associated with PD^4^.

To date, PD treatments are supportive and only provide symptom control; hence, there are still no curative treatments for PD that either inhibit or reverse the neurodegeneration. There is a need for new therapeutic agents acting on newly defined mechanisms, such as brain energy metabolism in PD, to overcome these translational failures. As reported in many neurodegenerative diseases, several lines of evidence have implicated bioenergy deficiency as a critical element in the pathogenesis of PD^12^. This is closely associated with mitochondrial failure and increased oxidative stress that eventually leads to neurodegeneration^13,14^. Relatedly, reduction of mitochondrial activity and downregulation of target genes involved in mitochondrial biogenesis has already been reported in patients with PD^15^. Furthermore, numerous reports have highlighted specific defects in mitochondrial, electron transport system genes, and protein components^16^. Previous multi-omics studies in humans have shown reduced chaperone proteins^17^ and dysregulated genes associated with mitochondrial energy metabolism such as complex I–IV, ATP synthase, and cytochrome C oxidase in PD^18,19^. Additionally, a very recent study evaluating the transcriptomic and proteomic levels of patients with PD suggested that PD should be considered as a disturbance of complex biologic systems^20^.

We hypothesized that PD patients could be treated with combined metabolic cofactor activators (CMA), including L-carnitine tartrate (to facilitate mitochondrial fatty acid uptake from the cytosol), nicotinamide riboside (the NAD^+^ precursor to induce neuronal mitochondrial β-oxidation and facilitate fatty acid transfer through the mitochondrial membrane), and the potent glutathione precursors L-serine and N-acetyl-l-cysteine (to reduce oxidative stress)^21–23^. We further hypothesized that supplementation with these metabolic cofactors would activate mitochondria and improve metabolism in brain. In animal toxicology studies and a human calibration study for CMA, we found that metabolic cofactors were well tolerated and increased the plasma levels of cofactors and their associated metabolites^24^. Additionally, supplementation with CMA effectively increased fatty acid oxidation and de novo glutathione generation, as judged using metabolomic and proteomic profiling^21^. In this placebo-controlled phase 2 study, we tested our hypotheses and the efficacy and safety of CMA in patients with PD.

## RESULTS

### CMA Improves Cognition and Blood Parameters in PD Patients

In the double-blind, randomized, placebo-controlled phase-2 study, we screened 65 PD patients and recruited 48 patients. We included patients older than 40 years with mild-to-moderate PD according to Hoehn Yahr scale 2 to 4. Of the 48 AD patients, 32 were randomly assigned to the CMA group and 16 to the placebo group (Figure 1A, Dataset S1). 5 patients dropped out of the study before Day 84 visit during the COVID-19 lockdown. On days 0, 28 and 84, we assessed the clinical variables and analyzed the differences between day 0 and day 28 as well as day 0 and day 84 in the CMA and placebo groups (Dataset S2).

**Figure 1.**
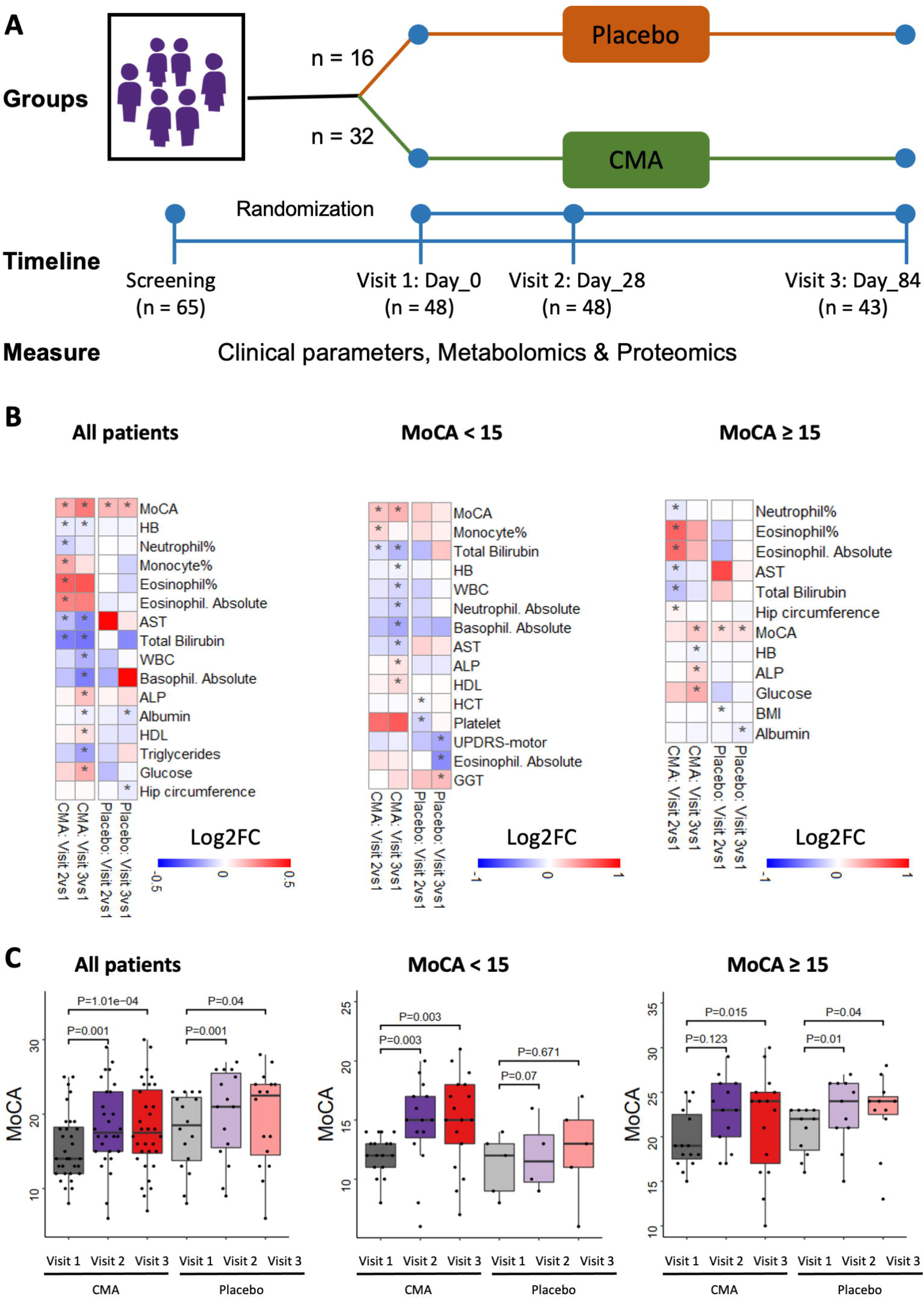
CMA Improves MoCA scores and clinical parameters. A) Study design for testing the effects of CMA in PD patients. B) Differences in MoCA scores in the CMA and placebo groups on Days 0, 28 and 84 are presented. Additionally, MoCA scores were analyzed by stratifying the patients into high- and low-scored MoCA groups (≥ 15 MoCA score is high, <15 is low). C) Heatmaps shows log2FC based alterations of the clinical variables compared to the CMA administration in both drug and placebo groups. Asterisks indicate statistical significance based on Student’s t-test. p-value <0.05. Log2FC: log2(fold change).

The mean age of the participants was 69.7 years (41–84 years), and 83.3% were men (Dataset S1). With regards to safety, no severe adverse events occurred, and 5 patients in the CMA group reported mild adverse events. All patients decided to complete the study. The safety profile of CMA in these patients was consistent with the results of our previous one-day calibration study^24^ and clinical trials^23,25^, including only a single component of CMA. Our present study showed that CMA was safe and well-tolerated in patients with PD.

We measured clinical variables in all patients and analyzed the differences before and after CMA administration in the active and placebo groups (Figure 1B and 1C, Dataset S1 and S2). As the increased Montreal Cognitive Assessment (MoCA) scores is known as the indicator of the increased cognitive functions, we observed that the mean MoCA scores were significantly higher in the CMA group both on Day 28 vs Day 0 (log2FoldChange(FC)=0.17, (13% improvement), p=0.001) and on Day 84 vs Day 0 (log2FC=0.27, (21% improvement), p= 0.0001). We also observed significant increased on MoCA scores in placebo group on Day 28 vs Day 0 (log2FC=0.16, (12% improvement), p=0.001) and on Day 84 vs day 0 (log2FC=0.15, (11% improvement), p=0.04) due to the recommendations of exercise and Mediterranean diet to all PD patients participated in the trial. Notably, the degree of increase of MoCA was much higher on Day 84 vs Day 0 in the CMA group than in the placebo group, suggesting the PD patients benefitted from CMA treatment after 84 days of treatment.

We also analyzed the differences of clinical parameters by stratifying the patients into high- and low-scored MoCA groups (≥ 15 MoCA score is high, <15 is low). Interestingly, we observed a significant improvement only in the low-scored patients in the CMA group both on Day 28 (log2FC=0.25, (19% improvement), p=0.003) and Day 84 (log2FC=0.32, (25% improvement), p=0.003), but no significance (p>0.05) was found in the low-scored placebo group (Figure 1B and 1C, Dataset S2). On the other hand, MoCA scores were significantly different in high-scored patients in the CMA (log2FC=0.22, (17% improvement), p=0.015) and placebo (log2FC=0.13, (9% improvement), p=0.04) groups Day 84 vs Day 0. These results suggest that both moderate patients (low-scored patients) and mild patients (high-scored patients) have a better response to CMA administration at Day 84 vs Day 0 compared to placebo.

Analysis of secondary outcome variables on Day 84 vs Day 0 showed that serum aspartate aminotransferase (AST) (log2FC= −0.23, p=0.03), total bilirubin (log2FC= −0.27, p=0.008) and triglycerides (log2FC= −0.17, p=0.04) levels were significantly lower in the CMA group (Figure 1B, Dataset S2). We also found significantly increased high-density lipoprotein (HDL) (log2FC= −0.07, p=0.04) only in the CMA group (Figure 1B, Dataset S2). In summary, we observed that the level of AST, total bilirubin, HDL and triglycerides were significantly improved due to the administration of CMA in PD patients as in the previously performed NAFLD Phase 2 clinical trial with CMA^26^.

We also measured the level of complete blood count parameters and found that their levels were significantly changed in the CMA group (Figure 1B, Dataset S2). We found that the levels of white blood cells (log2FC= −0.14, p=0.01) as well as the absolute number of basophile (log2FC= −0.25, p=0.04) were significantly lower in the CMA group on Day 84 vs Day 0. Of note, a decrease in the inflammatory parameters was also significant in the MoCA low-scored patients on Day 84 (Figure 1B, Dataset S2).

### Response to CMA Is Affected By Patients’ Clinical Profile

Variability in treatment response and clinical profiles among patients with PD is well documented^27^. As expected, we also observed this variability in our cohort. To harness this heterogeneity, we hypothesised that there exists some subset of patients, defined by clinical parameters, who would respond to CMA better than other patients.

We first determined whether alanine transferase (ALT), a marker for liver health status, could predict response to CMA. In order to do this, we stratified patients receiving CMA or placebo into high- and low-ALT groups and recorded MoCA with visit time (Figure 2A). We found that only the low ALT group exhibited increased MoCA score, but only when given CMA. No other group showed improvement to the same or better significance.

**Figure 2.**
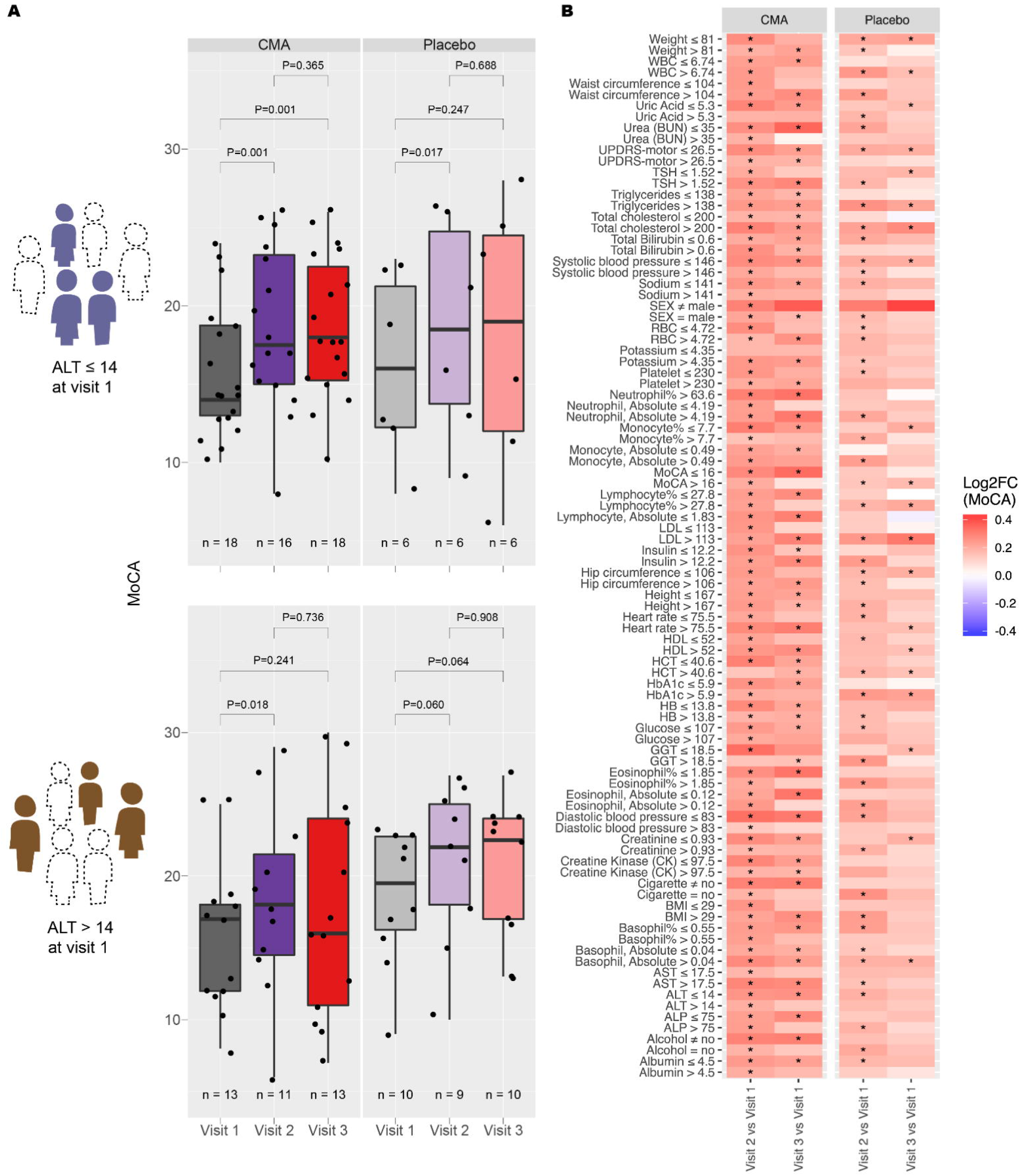
Identification of clinical measures informative for response to CMA. A) Distribution of MoCA scores over visit number for patients with ALT ≤ 14 at visit 1 (upper panel), and patients with ALT > 14 at visit 1 (lower panel). B) Between-visit changes to MoCA by clinical variable grouping. Only those groupings resulting in a more significant change to MoCA in CMA compared to placebo and with a p-value of 0.05 or better are shown. Colour scale indicates log2FC of MoCA between visits. Statistical significance between visits was determined by a paired t-test. *, p<0.05.

We then conducted the same stratification for each of the other clinical measures to determine other conditions in which CMA treatment leads to the best response (Figure 2B). In addition to the aforementioned low ALT, we found that low ALP, high AST, low GGT, low HCT, low glucose, low HbA1c, low uric acid, low eosinophil count, and low MoCA could indicate better response to CMA. This would indicate that CMA might work better in patients with underlying blood and glucose conditions.

### CMA Increases the Plasma Levels of Metabolites Associated with Metabolic Activators

We first analyzed the plasma levels of serine, carnitine, NR, and cysteine and found that administration of the CMA increased the plasma levels of serine, carnitine and nicotinamide proportionally on Day 84 vs Day 0 in the CMA group (Figure 3A, Dataset S3 & S4). In detail, the plasma levels of nicotinamide, nicotinurate, 1-methylnicotinamide, and N1-Methyl-2-pyridone-5-carboxamide (associated with NR and NAD^+^ metabolism); of serine, glycine and betaine (associated with serine and glycine metabolism); and of deoxycarnitine and carnitine (associated with carnitine metabolism) were significantly higher in the CMA group on Day 84.

**Figure 3.**
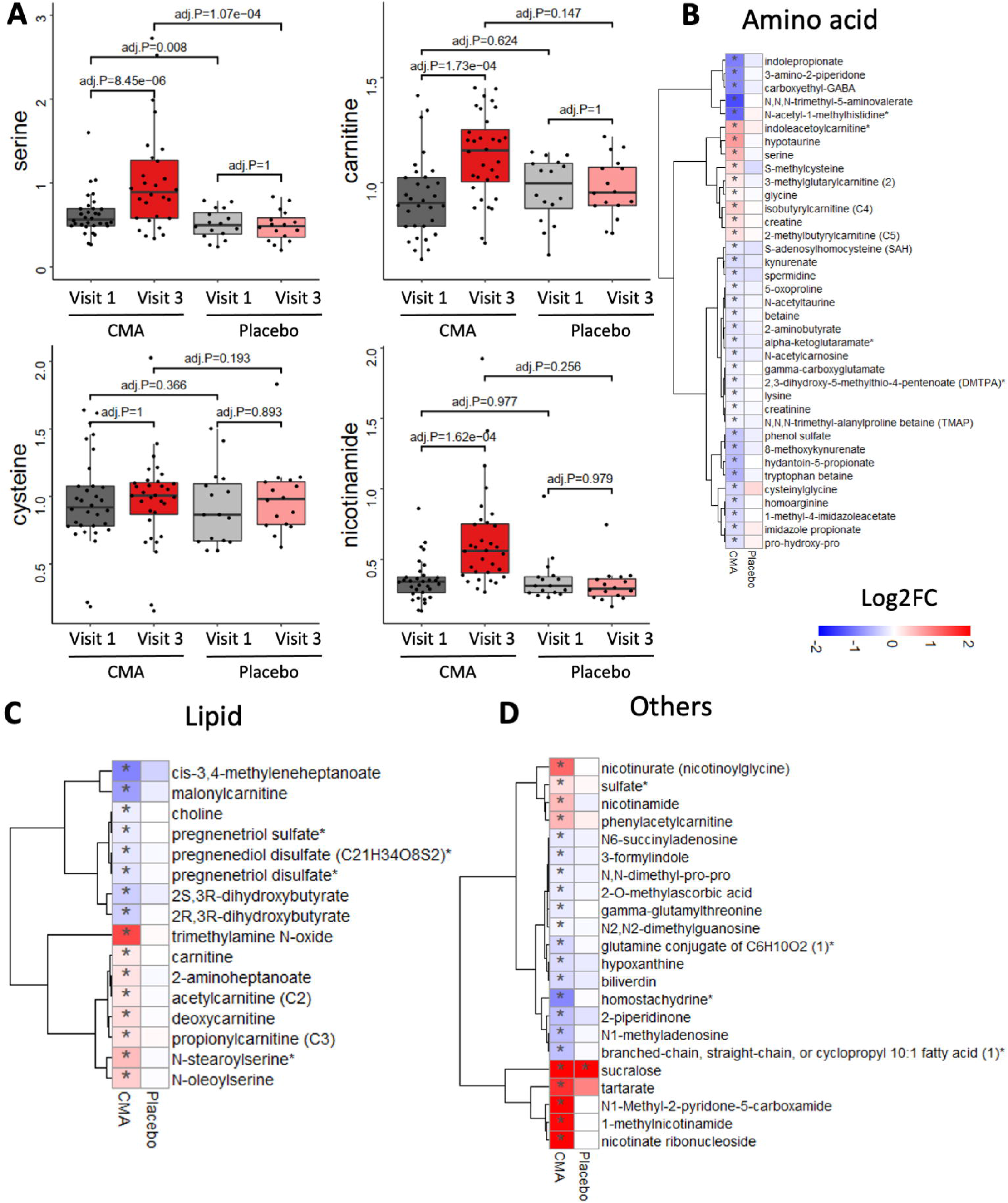
CMA alters plasma metabolite levels. A) Differences in the plasma levels of individual CMA, including serine, carnitine, cysteine and nicotinamide, are shown in the CMA and placebo groups on Days 0 and 84. Plasma levels of B) amino acids, C) lipids and D) other metabolites that are significantly different between Day 84 vs Day 0 in the CMA and placebo groups are presented. Adj. p< 0.05. Heatmap shows log2FC values of metabolites between Day 84 vs Day 0. Asterisks indicate statistical significance based on paired Student’s t-test. adj.p< 0.05. Log2FC: log2(fold change).

### Effect of CMA on Global Metabolism

We identified those plasma metabolites that were significantly (adj.p<0.05) different on Day 84 vs Day 0. We found that the plasma levels of 75 metabolites were significantly different in the CMA group (Figure 3, Dataset S4). Evaluation of plasma metabolites that differed significantly on Day 84 vs Day 0 in each group showed that the majority of metabolites related to amino acid (n=37) or lipid metabolism (n=16) and other metabolic pathways (n=22) were altered in the CMA group compared to the placebo group (Figure 3, Dataset S4).

Creatine is one of the most prevalent central nervous system (CNS) metabolites, and reduced levels have been associated with brain tissue injury^28^. Prior research has also shown that creatine is a central metabolite to maintain energy metabolism in brain^29,30^. In our study, we observed that plasma levels of creatine significantly increased on Day 84 vs Day 0 in the CMA group (Figure 3B, Dataset S4). Also upregulated on Day 84 vs Day 0 in the CMA group (Figure 3B, Dataset S4) was the metabolite glycine, which has been extensively investigated for its positive impact on cognitive performance^31,32^.

Elevated plasma homocysteine levels are known found to be associated with PD^33^ and several in-vivo studies have revealed the beneficial results of a low methionine diet on neurodegenerative diseases^33^. In our clinical trial, plasma levels of S-adenosylhomocysteine as well as 2,3-dihydroxy-5-methylthio-4-pentenoate and N-acetyl taurine were significantly decreased on Day 84 vs Day 0 in the CMA group (Figure 3B, Dataset S4).

Higher plasma concentrations of kynurenine pathway metabolites were related to CNS disorders^34^. In our study, we found that kynurenate, indolepropionate, 8-methoxy kynurenate and tryptophan betaine were significantly decreased on Day 84 vs Day 0 in the CMA group (Figure 3B, Dataset S4). Kynurenate is the product of tryptophan metabolism and is well known for its oxidative stress inducing effects by generating superoxide radicals and leading to cytochrome C depletion. According to the previous studies, high levels of kynurenine cause cell death in natural killer cells and decrease blood pressure in the systemic inflammatory response through reactive oxygen species^35,36^.

Accumulating evidence suggest an association between kidney and brain disorders, but the causal relationship between renal function and cognitive impairment remains to be established^37^. Recent studies showed that plasma levels of N,N,N-trimethyl-5-aminovalerate involved in lysine metabolism is an indicator of elevated urinary albumin excretion^38^. Here, we found that the plasma level of N,N,N-trimethyl-5-aminovalerate was significantly decreased on Day 84 vs Day 0 in the CMA group (Figure 3B, Dataset S4).

Moreover, the plasma level of creatinine was also significantly decreased on Day 84 vs Day 0 in the CMA group (Figure 3B, Dataset S4). Additionally, our analysis revealed reduced levels of several metabolites related to histidine metabolism in the CMA group on Day 84 vs Day 0. Among those N-acetyl-1-methylhistidine is related to decreased renal function.

Also, we found that plasma levels of metabolites related to the urea cycle (3-amino-2-piperidone, pro-hydroxy-pro, trimethyl-alanylproline betaine and homoarginine) were significantly lower in the CMA group on Day 84 vs Day 0 (Figure 3B, Dataset S4).

Lipids are central players in the pathogenesis of neurodegenerative diseases, including PD. Sphingolipids and cholesterol are not only structural components of plasma membranes but are also recognised as important components of brain function^39^. In our study, plasma levels of a considerable number of metabolites associated with carnitine and fatty acid metabolism were significantly elevated on Day 84 vs Day 0 in the CMA group (Figure 3C, Dataset S4). Of note, plasma levels of pregnenolone steroids and dihydroxy fatty acids were significantly decreased on Day 84 vs Day 0 (Figure 3C, Dataset S4).

Our additional analysis showed significantly upregulated carnitine metabolites, and significantly decreased plasma bilirubin metabolites, such as biliverdin (Figure 3D, Dataset S4). It has been shown to exert neuroprotective and procognitive effects in animal studies; this is especially appropriate given the altered levels found in our study. Carnitine, and NAD^+^ metabolites were shown to be involved in restoring the mitochondrial function, oxidative status and synaptogenesis^40^; while, in contrast, increased bilirubin and related metabolites led to neurotoxic effects in many PD models.

Despite its concentration-dependent dual antioxidant role^41^, studies on bilirubin in patients with PD have found increased levels of bilirubin^42–44^, possibly linked with the increased oxidative stress enzyme activity in patients with PD^44^. In support of this, there is evidence of increased hemoxygenase activity of dopaminergic cells after oxidative stress^45,46^, an enzyme responsible for the production of biliverdin^47^.

### Effect of CMA on Plasma Proteins

Plasma levels of 1466 protein markers were measured with the plasma proteome profiling platform Proximity Extension Assay quantifying the plasma level of target proteins. After quality control and exclusion of proteins with missing values in more than 50% of samples, 1463 proteins were analyzed (Dataset S5&S6). Proteins whose levels differed significantly between the visits in the CMA and placebo groups are listed in Dataset S6.

We analyzed the effect of CMA on plasma protein profile and found that 20 proteins were significantly (p <0.01) different in the CMA group on Day 84 vs Day 0. Thirteen of these proteins were significantly decreased, whereas seven of these proteins were significantly increased on Day 84 vs Day 0. Among these proteins, we found that the plasma levels of OSM, MMP9, RASSF2, GSTP1, GZMH, FEN1, NCF2, MNDA, AK1, AZU1, AARSD1 and RAPGAP1L were significantly downregulated. The plasma levels of KLB, GPA33, SLC39A14, IL17RB, LRIG1, ALPP, and SERPINB5 were significantly upregulated only in the CMA group (Figure 4A & Dataset S6). We observed that IL1B, CXCL6, TPT1, PIK3AP, ARHGAP1, CXCL11, PPME1, AKT1S1 and RILP were significantly (p<0.01) downregulated and KLB upregulated in the placebo group (Figure 4A, Dataset S6). Several experimental studies suggest that the altered proteins found in our study are involved in inflammation, membrane transport, DNA repair, membrane trafficking, synaptogenesis, oxidative injury, and protein aggregation. For instance, ALPP and LRIG1 are well-known molecules for their antioxidant and neurotrophic properties among the increased proteins. Similarly, SLC39A14 functions as a pivotal manganese transporter in vertebrates^48^ and its deficiency is associated with rapidly progressive childhood-onset parkinsonism–dystonia due to excessive accumulation of manganese in the brain. Also, GPA33, a protein strictly limited to the intestine and responsible for intestinal integrity with unknown central functions^49^, was found to be increased, which might suggest the role of a gut-brain axis component in neurodegenerative disorders.

**Figure 4.**
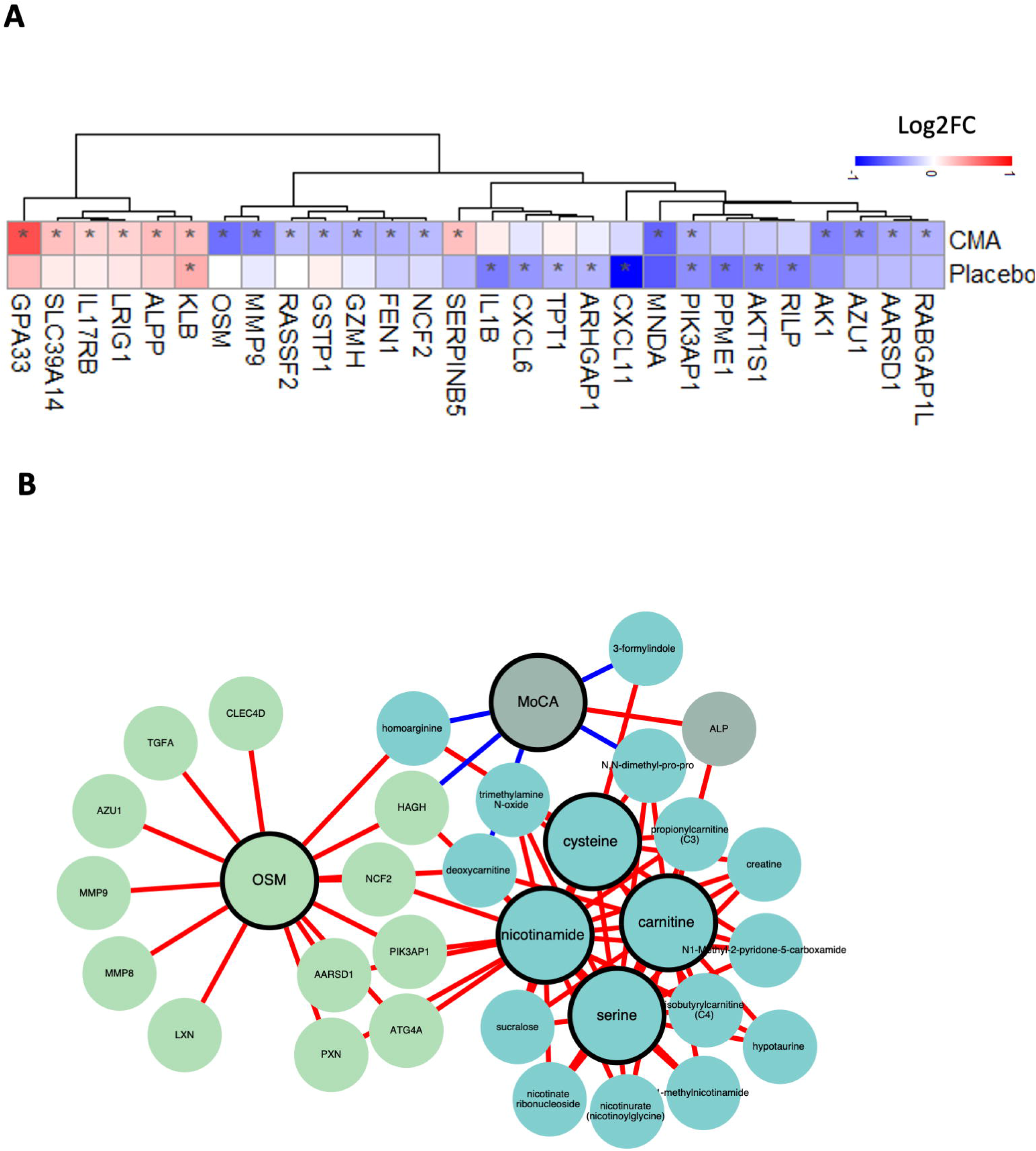
Altered plasma protein levels and integrated multi-omics network. A) Heatmap shows log2FC based alterations between the significantly different proteins on Day 84 vs Day 0 in the CMA and placebo groups. Asterisks indicate statistical significance based on paired Student’s t-test. p < 0.01. B) Integrated multi-omics data based on network analysis represents the neighbours of the CMA, including serine, carnitine, nicotinamide and cysteine, and MoCA scores. Only analytes that are significantly altered in CMA Day 84 vs Day 0 are highlighted.

Similar relevant alterations were also observed for decreased protein levels. For instance, AK1, and ATP regulator protein, has been defined in postmortem PD brains as upregulated, indicating energy dysregulation in PD^50^, which is especially relevant considering the increasing evidence of a strong link between protein aggregation, inflammation and energy deficiency in PD. We consistently found significantly reduced levels of MMP9, RASSF2, GSTP1, GNZMH, NCF-2, AARSD1, MNDA, OSM, and FEN1 which are proteins involved in neuroinflammation, apoptosis, oxidative stress, central and even peripheral immunologic responses.

Other notable observations include the down-regulated levels of OSM, MNDA, AZU1 and MMP9, which are well known neuroinflammatory markers, proven in several PD models. Similar beneficial alterations have also been observed for some other proteins such as RABGAP1L and single tRNA synthetase editing domain, involved directly or indirectly in misfolded protein aggregation. It should be noted that we observed changes in some other molecules that are also consistent with human data, showing either a significant improving effect on the neurodegenerative process of being involved in the pathogenetic process. For instance, AZU1 and MMP-9, both important cascades of a multifunctional neuroinflammatory process and blood-brain barrier breakdown, as mentioned above, were elevated in individuals with PD^51^. Also, dysregulated levels of RABGAP1L, involved in cellular membrane trafficking, and GSTP1, a well-known molecule with attenuating functions on oxidative and endoplasmic reticulum stress^52,53^, have been found in human dopaminergic neurons and the synaptosomal fraction of patients with PD^54^. In addition, LRIG1, was increased in the present study, and was recently shown to be located in the soma and extends out into the apical dendrites of hippocampal pyramidal neurons, controlling brain-derived neurotrophic factor signalling^55^, which is a neuroprotective and pro-cognitive molecule.

### Integrative Multi-Omics Analysis

The integrations of multi-omics data have been previously shown to be beneficial in understanding diseases^56^. We generated a PD-specific network based on multi-omics (metabolomics and proteomics) data, complemented by clinical chemistry and anthropometrics data, generated in this study. The main goal of the network analysis was to elucidate the functional relationships between analytes within and between different omics and data types. The network was generated using the same pipeline as iNetModels^57^, an interactive multi-omics network database and visualization tool, where we deposited the full network from this study. The generated network has ∼2 million edges from 2295 nodes (40% network density, Dataset S7).

To understand the CMA and MoCA interactions, we extracted a subnetwork of those analytes and their top neighbours (Figure 4B). From the subnetwork, we observed that MoCA was associated with the plasma levels of serine, trimethylamine N-oxide (phospholipid), deoxycarnitine, creatine, and several nicotinate and nicotinamide metabolites (1-methylnicotinamide, nicotinurate, nicotinate ribonucleoside, and N1-methyl-2-pyridone-5-carboxamide). MoCA was also associated with the plasma levels of several proteins, including MMP9 and OSM, highlighted in the previous section. The same metabolites and proteins were shown to be positively correlated to the CMA.

We also performed a centrality analysis to identify the critical nodes in the network. The top 10 most central metabolites were dominated by xenobiotics metabolites, including those associated with neurological and psychoactive drugs (lamotrigine, O-desmethylvenlafaxine, venlafaxine, and diazepam). We also observe a nicotinamide-related metabolite (Adenosine diphosphate (ADP)-ribose) in that list. Meanwhile, the top 10 proteins included nervous system-related protein (NPY), a regulator of the TGF-beta process (ITGB6), and T-cell activation regulator (PRKAR1A). These results showed that integrative multi-omics network analysis may elucidate the functional relationships between analytes, support the results from single omics data and add new insights by discovering key analytes from the network.

### Common Effect of CMA on Plasma Metabolites and Proteins in AD And PD Patients

We validated our findings in an independent AD cohort (n=60) who received the same protocol of CMA therapy and showed a significant improvement in cognitive functions^25^. Comparison of metabolomics data in the treated groups of both AD and PD cohorts than their placebo on Day 84 vs Day 0 revealed that the plasma levels of 60 metabolites were significantly altered in the same direction (Figure 5, Dataset S8). These changes were in amino acid (n=30), lipid metabolism (n=14) and other metabolic pathways (n=16) (Figure 5, Dataset S8).

**Figure 5.**
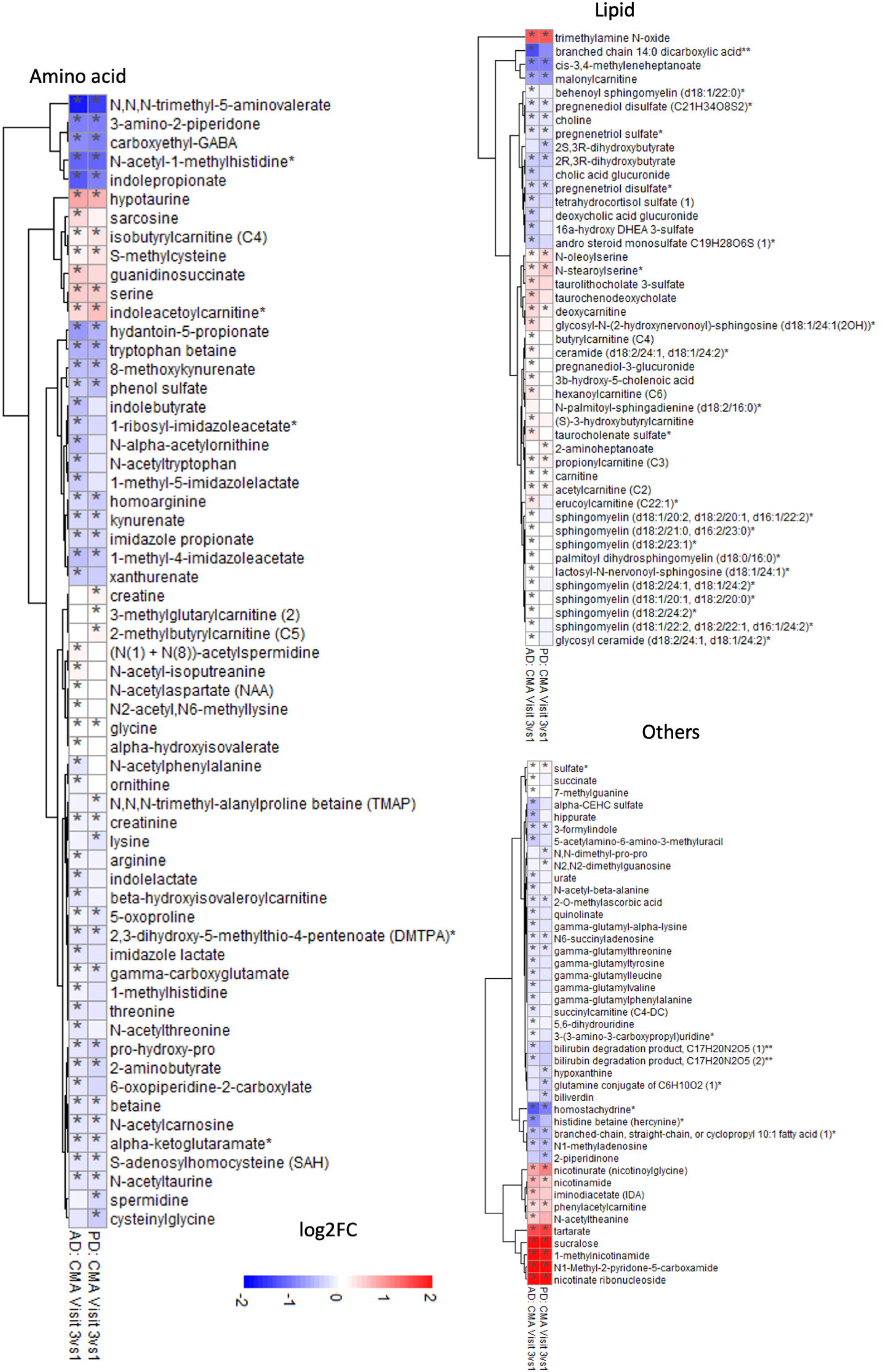
Common significantly altered metabolites in PD and AD clinical trials. Plasma levels of A) amino acids, B) lipids and C) other metabolites that are significantly different between Day 84 vs Day 0 in the CMA group of this study and an independent AD trial^25^ are presented. Adj. p< 0.05. Heatmap shows log2FC values of metabolites between Day 84 vs Day 0. Asterisks indicate statistical significance based on paired Student’s t-test. adj.p< 0.05. Log2FC: log2(fold change).

Primarly, the plasma levels of CMA constituents and byproducts were increased in the CMA groups of both cohorts (Figure 5A, Dataset S8). Additionally, hypotaurine and trimethylamine N-oxide were also significantly increased in both CMA groups (Figure 5A, Dataset S8). On the other hand, plasma levels of numerous amino acids in key pathways (i.e glutamate, glutathione, histidine and tryptophan metabolism and urea cycle) were significantly decreased in the both CMA groups (Figure 5A, Dataset S8). Also, creatinine, betaine, N,N,N-trimethyl-5-aminovalerate and phenol sulfate plasma levels were significantly downregulated in CMA groups than placebo (Figure 5A Dataset S8). Furthermore, plasma levels of metabolites associated with fatty acids, choline and pregnenolone steroids were significantly decreased in the CMA groups of both cohorts (Figure 5B, Dataset S8). Interestingly, plasma levels of 2-O-methylascorbic acid and metabolites in purine metabolism were also significantly decreased in the both CMA groups (Figure 5C, Dataset S8).

We also compared the plasma proteomic profile between two cohorts and found that a significant increse in the plasma level of KLB and a significant decrease in the plasma level of OSM in both on Day 84 vs Day 0 in the CMA groups (Dataset S8). Our results collectively indicated that CMA improved cognitive functions in different patients groups by similar alterations on plasma profiles.

## DISCUSSION

In our study, we observed that CMA treatment significantly improved cognitive function in patients with PD based on MoCA scores. We found that the cognitive functions in PD patients is improved 21% in the CMA group, whereas it is improved only 11% in the placebo group after 84 days of CMA administration. Our finding were in agreement with the results of our recently performed AD clinical phase 2 study, where we administed CMA to 60 AD patients using the same protocol ^25^. In the AD study, we assessed the cognitive functions with AD Assessment Scale-cognitive subscale (ADAS-Cog) score and found that the ADAS-Cog scores is improved 29% in the CMA group whereas it is improved only 14% in the placebo group after 84 days of CMA administration. Around 11-14% improvement in the placebo group in both PD and AD clinical trials can be explained with the recommendations of exercise and Mediterranean diet to all participated patients.

In evaluating the metabolomic parameters, we found that levels of plasma nicotinamide, carnitine, cysteine, and serine were significantly elevated. Considering the role of these metabolites in the pathogenesis of PD, it was not surprising to see such beneficial alterations after CMA treatment. For instance, NAD^+^ has been shown to restore bioenergy imbalance in animal PD models and patients with PD, offering significant alleviation of motor symptoms correlated with the increase of L-dopa availability^58,59^. A similar situation also applies to other metabolites such as serine, cysteine, and carnitine.

Beyond their critical role in mitochondrial energy deficiency confirmed by many PD models, there are a growing number of studies suggesting that these metabolites are also involved in patients with PD. For instance, among many metabolites, plasma serine and carnitine levels have been shown to have the strongest inverse correlation with PD severity^60^. These clinical data accorded well with their beneficial role in oxidative injury^61^ (increasing the synthesis of glutathione) and impaired mitochondrial bioenergetics in PD^62^. Also, a down-regulation of carnitine and its metabolites have been shown in patients with PD^63^. As for NAD^+^, recent human data suggest that it is also linked to improved behavioural and motor symptoms in patients with PD^64^, possibly mediated by the glutamatergic role of serine^65^ when applied either separately or in combination.

Several animal and human studies provided strong evidence for countering some of the effects of disturbed carnitine metabolism with acetyl-L-carnitine^66,67^. Here, it is worth mentioning that acetyl-L-carnitine treatment seems to induce both cerebral energy metabolism and cholinergic neurotransmission^68^, which may have also led to improved cognition in our patients with PD. These findings suggest that in addition to increased carnitine levels, decreased choline metabolites may also have mediated the pro-cognitive effect of our treatment. Thus, our finding of improved lipid metabolites is of critical significance, as altered lipid metabolism and cognitive dysfunction are both known to occur during the neurodegenerative process.

Similar beneficial alterations were also observed in dihydroxybutyrate (DHBA) levels. Our finding of decreased DHBA levels might indicate the restoration of alternative energy production pathways, such as GABA shunt^69^, activated during the cellular energy composition, as in AD^69^ and PD^70^. Confirmingly, in our recent AD study, we found similarly decreased post-therapeutic levels of DHBA^25^, suggesting an energy deficiency seen in both neurodegenerative diseases, but also the pro-energy role of CME. Beyond their role in energy mechanisms, lipids are involved in many critical intracellular signaling and transporting processes, as the main component of cellular membranes, which make them a strong candidate for cognition, even in healthy individuals^71^. However, under neurodegenerative conditions characterized by disturbed lipid metabolism, their behavior shifts to become more pro-inflammatory and oxidative, rather than regulatory, and contributes significantly to the acceleration of the neurodegenerative process, as in PD^72^. A good example is the link between lipid dysregulation and alpha-synuclein aggregation, leading to dopaminergic neuronal death in PD^72^.

Our analysis also revealed decreased purine metabolism in the CMA group. Plasma levels of hypoxanthine and adenosine were significantly reduced in the CMA group, consistent with a recent clinical study showing increased hypoxanthine levels in PD^73^. Furthermore, our findings of decreased peripheral levels of hypoxanthine could also be interpreted as increased CNS bioavailability of hypoxanthine considering its high blood-brain barrier permeability and adenosine triphosphate (ATP)-enhancing role under energy crisis conditions.

Considering that creatine is an essential part of mitochondrial therapy for many neurodegenerative diseases^74^, and exerts significant neuroprotection in several in-vitro and in-vivo PD studies^75^, our findings of increased creatine levels were not surprising to us. Consistently, creatine supplementation improves cognition not only in healthy individuals^75^, but also in PD patients with cognitive impairment when combined with another mitochondrial energetic molecule, coenzyme q10^76^. Also, its combination with minocycline significantly is known to reduce PD progression in early stages^77^.

Human data have shown that creatine exerted its pro-cognitive effect through increasing the brain oxygen utilization^78^ fitting well with its pro-energetic role. We also have found decreased creatinine levels, compatible with recent PD clinical data showing an inverse correlation between uric acid/creatinine ratios and PD progression^79^. Similarly, increased serum creatinine was related to incident dementia and cognitive impairment^80^.

Moreover, we found that the plasma level of 3-amino-2-piperidone associated with urea, especially in ornithine metabolism, was significantly decreased after CMA. Although it remains unclear how decreased urea metabolism is associated with our treatment effect, several studies have indicated that urea pathways might be perturbed in PD^81–83^, indicating increased ornithine levels in PD. This is consistent with the detrimental role of high urea on learning^84^.

We found decreased levels of homostachydrine, categorized under the xenobiotics class, which are generally not endogenously synthesized but can be produced by the gut microbiome^85^. Although no biological role has been defined for homostachydrine, a recent study reported that it was involved in the increased inflammatory process in an experimental autoimmune encephalomyelitis model in mice^86^. This observation agrees well with the identified decreased levels of homostachydrine after CMA treatment.

It is worth mentioning here that hypotaurine metabolism plays a vital role in fighting against neurodegeneration through the production of secondary bile acids, which exert a significant neuroprotective effect^87^. A recent study identified increased hypotaurine metabolism as a compensatory neuroprotective pathway in a mouse model with alpha-synuclein^88^. Also, N-stearyl serine, a lipoamino acid, with many pleiotropic signalling functions^89^, was recently shown to be positively associated with cognition in patients with AD, especially those with good adherence to a Mediterranean diet^90^. Thus, stearoyl-CoA desaturase, a rate-limiting enzyme in the biosynthesis of monounsaturated fatty acids, has been suggested as a new player mediating lipid metabolism with synuclein aggregation^90^.

It is difficult to explain why we observed only an improvement in cognitive scores but no improvement in Unified Parkinson Disease Rating Scale (UPDRS) motor scores; despite a robust cognitive response, we observed no significant alteration in motor scores (Dataset S2). This is, however, in line with previous studies showing that energy metabolic supplementations failed to show a clear benefit in PD^91^, suggesting that disturbed energy metabolism cannot be easily reversed in already dead circuits. Early stages of PD are likely to be responsive to such metabolic approaches. For instance, besides some critical differences in affected regions between patients with PD-with and without cognitive impairment^91,92^-the metabolic expression of both clinical patterns also increases at different rates. PD studies have suggested that cognitive expression has a slower metabolic deterioration rate than the pure motor pattern^93^, which would indicate probable later neurodegeneration, and explain the selective therapeutic response observed in our study.

Also, our improved MoCA scores, along with increased post-therapeutic alterations in lipid metabolites, fits well with recent metabolomics PD studies showing an inverse correlation between lipid metabolites and MoCA scores^94^, suggesting the role of lipid metabolite alterations is a key parameter in discriminating PD with cognitive impairment from regular PD patients.

A few limitations of the study need to be considered. First, we evaluated the treatment effect using only omics-based methods and clinical evaluation without neuroimaging. Thus, a clinical trial combined with neuroimaging methods to delineate the effects of CMA on functional and structural brain alterations would be informative. Second, the link between systemic and CNS alterations and their relations to the central protein aggregations defined as critical in PD has not been evaluated and further amyloid or tau-based neuroimaging combined with CSF evaluations can be planned.

To date, there is no curative medical treatment for cognitive impairment in PD; therapeutic options are limited to cholinesterase inhibitors^95^, which may be transient to replace the impaired cholinergic transmission^96^. Also, it is still under debate whether dopamine replacement medications have cognitive side effects^97^ while improving motor symptoms in cognitively impaired PD patients. To that end, there is still no available drug able to revert PD cognitive deficits, and, unfortunately, current treatment approaches available for motor impairment are not devoid of cognitive side effects. Considering all of these findings, it is crucial to better delineate the neuropathology underlying cognitive symptoms through clinical and preclinical studies. In conclusion, CMA significantly improved cognition and serum markers of PD after 84 days. These findings suggest that targeting multiple pathways using CMA is a potentially effective therapeutic strategy for PD as previously shown in AD.

## MATERIAL AND METHODS

### Trial Design and Oversight

Patients for this randomized, double-blinded, placebo-controlled, phase-2 study were recruited at the Faculty of Medicine, Alanya Alaaddin Keykubat University, Antalya, Turkey and Faculty of Medicine, Istanbul Medipol University, Istanbul, Turkey. Written informed consent was obtained from all participants before the initiation of any trial-related procedures. An independent external data-monitoring committee oversaw the safety of the participants and the risk-benefit analysis. The trial was conducted in accordance with Good Clinical Practice guidelines and the principles of the Declaration of Helsinki. The study was approved by the ethics committee of Istanbul Medipol University, Istanbul, Turkey, and retrospectively registered at https://clinicaltrials.gov/ with Clinical Trial ID: NCT04044131.

### Participants

Patients were enrolled in the trial if they were over 40 years of age with mild to moderate PD according to Hoehn Yahr scale 2 to 4. Patients who had a history of stroke, severe brain trauma, toxic drug exposure were excluded. Also, patients who indicate to Parkinson-Plus syndrome (i.e., pyramidal, cerebellar and autonomic dysfunction findings and gaze paralysis) in the neurological examination were omitted. The inclusion, exclusion, and randomization criteria are described in detail in the Supplementary Appendix.

### Randomization, Interventions, and Follow-up

Patients were randomly assigned to receive CMA or placebo (2:1). Patient information (patient number, date of birth, initials) was entered into the web-based randomization system, and the randomization codes were entered into the electronic case report form. All clinical staff were blinded to treatment, as were the participants.

Treatment started on the day of diagnosis. Both placebo and CMA were provided in powdered form in identical plastic bottles containing a single dose to be dissolved in water and taken orally one dose in the morning after breakfast and one dose in the evening after dinner. Each dosage of CMA dose contained a 3.73 g L-carnitine tartrate, 2.55 g N-acetylcysteine, 1 g nicotinamide riboside chloride, and 12.35 g serine. All patients returned for a follow-up visit on Day 84. Further information is provided in the Supplementary Appendix.

### Outcomes

The primary endpoint in the original protocol was to assess the clinical efficacy of CMA in PD patients. For the primary purpose, the clinical differences in cognition of subjects receiving twelve-week treatment either with metabolic cofactors supplementation or placebo were determined. The primary analysis was on the difference in cognitive and motor function scores between the placebo and the treatment arms. The motor, cognitive and behavioural functions of PD patients were evaluated via Unified Parkinson Disease Rating Scale (UPDRS), The Montreal Cognitive Assessment (MoCA) and Neuropsychiatric Inventory (NPI), respectively. The secondary aim of this study was to evaluate the safety and tolerability of CMA. All protocol amendments were authorized and approved by the sponsor, the institutional review board or independent ethics committee, and the pertinent regulatory authorities.

The number and characteristics of adverse events, serious adverse events, and treatment discontinuation due to CMA were reported from the beginning of the study to the end of the follow-up period as key safety endpoints. The changes in vital signs, baseline values, and the status of treatment were recorded on day 0 and 84. A complete list of the endpoints is provided in the Supplementary Appendix.

### Proteomics Analysis

Plasma levels of proteins were determined with the Olink panel (Olink Bioscience, Uppsala, Sweden). Briefly, each sample was incubated with DNA-labeled antibody pairs (proximity probes). When an antibody pair binds to its corresponding antigens, the corresponding DNA tails form an amplicon by proximity extension, which can be quantified by high-throughput, real-time PCR. Probe solution (3 μl) was mixed with 1 μl of sample and incubated overnight at 4°C. Then 96 μl of extension solution containing extension enzyme and PCR reagents for the pre-amplification step was added. The extension products were mixed with detection reagents and primers and loaded on the chip for qPCR analysis with the BioMark HD System (Fluidigm Corporation, South San Francisco, CA). To minimize inter- and intrarun variation, the data were normalized to both an internal control and an interplate control. Normalized data were expressed in arbitrary units (Normalized Protein eXpression, NPX) on a log2 scale and linearized with the formula 2NPX. A high NPX indicates a high protein concentration. The limit of detection, determined for each of the assays, was defined as three standard deviations above the negative control (background).

### Untargeted Metabolomics Analysis

Plasma samples were collected on Days 0 and 84 for nontargeted metabolite profiling by Metabolon (Durham, NC). The samples were prepared with an automated system (MicroLab STAR, Hamilton Company, Reno, NV). For quality control purposes, a recovery standard was added before the first step of the extraction. To remove protein and dissociated small molecules bound to protein or trapped in the precipitated protein matrix, and to recover chemically diverse metabolites, proteins were precipitated with methanol under vigorous shaking for 2 min (Glen Mills GenoGrinder 2000) and centrifuged. The resulting extract was divided into four fractions: one each for analysis by ultraperformance liquid chromatography– tandem mass spectroscopy (UPLC-MS/MS) with positive ion-mode electrospray ionization, UPLC-MS/MS with negative ion-mode electrospray ionization, and gas chromatography– mass spectrometry; one fraction was reserved as a backup.

### Statistical Analysis

Paired t-tests were used to identify the differences in clinical parameters between time points, and one-way ANOVA was used to find the shifts between CMA and placebo groups at each time point. We removed the metabolite profiles with more than 50% missing values across all samples to analyse plasma metabolomics. Metabolite changes between time points were analyzed by paired t-test. One-way ANOVA analyzed metabolite changes between CMA and placebo groups. Missing values were removed in pairwise comparison. The p-values were adjusted by Benjamini & Hochberg method. Metabolites with a false-discovery rate of 5% were considered statistically significant.

We removed the protein profiles with more than 50% missing values across all samples to analyse plasma proteomics. A paired t-test was used to identify the changes between time points, and one-way ANOVA was used to determine the changes between different groups. p<0.01 was considered statistically significant.

For the identification of clinical variables that inform response to CMA, low-scored and high-scored patient groups were established for each clinical variable on the basis of the median score at day 0. Patients scoring equal to or less than the median score were assigned “low”; patients scoring greater than the median score were assigned “high”. Statistical significance in the difference between MoCA score distributions over visit number was tested between visits using paired t-tests. Clinical variables were deemed informative to predict the response to CMA if exactly one group, low or high, showed more statistically significant changes in MoCA in the CMA group than in the placebo group.

### Generation of Multi-Omics Network

A Multi-omics network was generated based on the Spearman correlations, and the significant associations (adj.p < 0.05) are presented. The analyses were performed with the SciPy package in Python 3.7. Centrality analysis on the network was performed using iGraph Python.

## Supporting information

Yulug_PD_Dataset S1

Yulug_PD_Dataset S2

Yulug_PD_Dataset S3

Yulug_PD_Dataset S4

Yulug_PD_Dataset S5

Yulug_PD_Dataset S6

Yulug_PD_Dataset S7

Yulug_PD_Dataset S8

## Data Availability

All available data included in the supplementary datasets

## ACKNOWLEDGMENTS

This work was financially supported by ScandiBio Therapeutics and Knut and Alice Wallenberg Foundation. The authors would like to thank Metabolon Inc. (Durham, USA) for generating metabolomics data and ChromaDex Inc. (Irvine, CA, USA) for providing NR. AM and HY acknowledge support from the PoLiMeR Innovative Training Network (Marie Skłodowska-Curie Grant Agreement No. 812616), which received funding from the European Union’s Horizon 2020 research and innovation programme.

The computations and data handling were enabled by resources provided by the Swedish National Infrastructure for Computing (SNIC) at UPPMAX, partially funded by the Swedish Research Council through grant agreement no. 2018-05973.

## CONFLICT OF INTEREST

AM, JB and MU are the founder and shareholders of ScandiBio Therapeutics. The other authors declare no competing interests.

## SUPPORTING INFORMATION

Supporting Information includes 8 Supplementary Datasets.

## SUPPLEMENTARY DATASET LEGENDS

**Dataset S1**. Collection of samples of CMA and placebo groups and the measured values of clinical indicators before and after treatment.

**Dataset S2**. Statistical analysis of clinical indicators between different visits or groups.

**Dataset S3**. Plasma metabolomics data for each patient before and after treatment.

**Dataset S4**. Statistical analysis of plasma metabolites between different visits or groups.

**Dataset S5**. Plasma proteomics data was generated with the Olink cardiometabolic, inflammation, neurology and oncology panels for each patient before and after treatment.

**Dataset S6**. Statistical analysis of plasma proteins between different visits or groups.

**Dataset S7**. Multi-Omics Network Data, including edges and nodes information. The network is presented in the iNetModels (http://inetmodels.com).

**Dataset S8**. Common significant metabolites and proteins in this study and in an independent Alzheimer’s study^25^

## Notes

### Clinical Trial

NCT04044131

### Author Declarations

The study was approved by the ethics committee of Istanbul Medipol University, Istanbul, Turkey

## REFERENCES

1. Spillantini, M.G., Crowther, R.A., Jakes, R., Hasegawa, M. & Goedert, M. alpha-Synuclein in filamentous inclusions of Lewy bodies from Parkinson’s disease and dementia with lewy bodies. Proceedings of the National Academy of Sciences of the United States of America 95, 6469–6473 (1998).

2. Feigin, V.L., et al. Global, regional, and national burden of neurological disorders during 1990–2015: a systematic analysis for the Global Burden of Disease Study 2015. The Lancet Neurology 16, 877–897 (2017).

3. Dorsey, E.R. & Bloem, B.R. The Parkinson Pandemic-A Call to Action. JAMA Neurol 75, 9–10 (2018).

4. Chaudhuri, K.R. & Sauerbier, A. Parkinson disease. Unravelling the nonmotor mysteries of Parkinson disease. Nat Rev Neurol 12, 10–11 (2016).

5. Bosco, D., et al. Dementia is associated with insulin resistance in patients with Parkinson’s disease. J Neurol Sci 315, 39–43 (2012).

6. Pappatà, S., et al. Mild cognitive impairment in drug-naive patients with PD is associated with cerebral hypometabolism. Neurology 77, 1357–1362 (2011).

7. Camargo Maluf, F., Feder, D. & Alves de Siqueira Carvalho, A. Analysis of the Relationship between Type II Diabetes Mellitus and Parkinson’s Disease: A Systematic Review. Parkinsons Dis 2019, 4951379 (2019).

8. Krikorian, R., et al. Nutritional ketosis for mild cognitive impairment in Parkinson’s disease: A controlled pilot trial. Clinical Parkinsonism & Related Disorders 1, 41–47 (2019).

9. Bohnen, N.I., et al. Cerebral glucose metabolic features of Parkinson disease and incident dementia: longitudinal study. J Nucl Med 52, 848–855 (2011).

10. Brakedal, B., et al. Glitazone use associated with reduced risk of Parkinson’s disease. Mov Disord 32, 1594–1599 (2017).

11. Athauda, D., et al. Exenatide once weekly versus placebo in Parkinson’s disease: a randomised, double-blind, placebo-controlled trial. Lancet 390, 1664–1675 (2017).

12. González-Casacuberta, I., Juárez-Flores, D.L., Morén, C. & Garrabou, G. Bioenergetics and Autophagic Imbalance in Patients-Derived Cell Models of Parkinson Disease Supports Systemic Dysfunction in Neurodegeneration. Front Neurosci 13, 894 (2019).

13. Schapira, A.H.V. & Gegg, M. Mitochondrial contribution to Parkinson’s disease pathogenesis. Parkinsons Dis 2011, 159160–159160 (2011).

14. Lin, M.T. & Beal, M.F. Mitochondrial dysfunction and oxidative stress in neurodegenerative diseases. Nature 443, 787–795 (2006).

15. Siddiqui, A., et al. Mitochondrial Quality Control via the PGC1α-TFEB Signaling Pathway Is Compromised by Parkin Q311X Mutation But Independently Restored by Rapamycin. The Journal of Neuroscience 35, 12833 (2015).

16. Saxena, U. Bioenergetics failure in neurodegenerative diseases: back to the future. Expert Opin Ther Targets 16, 351–354 (2012).

17. Caudle, W.M., Bammler, T.K., Lin, Y., Pan, S. & Zhang, J. Using ‘omics’ to define pathogenesis and biomarkers of Parkinson’s disease. Expert Rev Neurother 10, 925–942 (2010).

18. Miller, R.M., et al. Robust dysregulation of gene expression in substantia nigra and striatum in Parkinson’s disease. Neurobiol Dis 21, 305–313 (2006).

19. Duke, D.C., et al. Transcriptome analysis reveals link between proteasomal and mitochondrial pathways in Parkinson’s disease. Neurogenetics 7, 139–148 (2006).

20. Ruffini, N., Klingenberg, S., Schweiger, S. & Gerber, S. Common Factors in Neurodegeneration: A Meta-Study Revealing Shared Patterns on a Multi-Omics Scale. Cells 9(2020).

21. Mardinoglu, A., et al. An Integrated Understanding of the Rapid Metabolic Benefits of a Carbohydrate-Restricted Diet on Hepatic Steatosis in Humans. Cell Metab 27, 559–571.e555 (2018).

22. Mardinoglu, A., et al. The Potential Use of Metabolic Cofactors in Treatment of NAFLD. Nutrients 11, 1578 (2019).

23. Altay, O., et al. Combined Metabolic Activators Accelerates Recovery in Mild-to-Moderate COVID-19. Advanced Science n/a, 2101222 (2021).

24. Zhang, C., et al. The acute effect of metabolic cofactor supplementation: a potential therapeutic strategy against non-alcoholic fatty liver disease. Mol Syst Biol 16, e9495 (2020).

25. Yulug, B., et al. Combined Metabolic Activators Improves Cognitive Functions in Alzheimer’s Disease. medRxiv, 2021.2007.2014.21260511 (2021).

26. Zeybel, M., et al. Combined Metabolic Activators Reduces Liver Fat in Nonalcoholic Fatty Liver Disease Patients. medRxiv, 2021.2005.2020.21257480 (2021).

27. Greenland, J.C., Williams-Gray, C.H. & Barker, R.A. The clinical heterogeneity of Parkinson’s disease and its therapeutic = implications. Eur J Neurosci 49, 328–338 (2019).

28. Béard, E. & Braissant, O. Synthesis and transport of creatine in the CNS: importance for cerebral functions. Journal of Neurochemistry 115, 297–313 (2010).

29. Bonilla, D.A., et al. Metabolic Basis of Creatine in Health and Disease: A Bioinformatics-Assisted Review. Nutrients 13, 1238 (2021).

30. Tachikawa, M., Hosoya, K., Ohtsuki, S. & Terasaki, T. A novel relationship between creatine transport at the blood-brain and blood-retinal barriers, creatine biosynthesis, and its use for brain and retinal energy homeostasis. Subcell Biochem 46, 83–98 (2007).

31. de Bartolomeis, A., et al. Glycine Signaling in the Framework of Dopamine-Glutamate Interaction and Postsynaptic Density. Implications for Treatment-Resistant Schizophrenia. Front Psychiatry 11, 369–369 (2020).

32. Avila, A., Nguyen, L. & Rigo, J.-M. Glycine receptors and brain development. Frontiers in cellular neuroscience 7, 184–184 (2013).

33. Fan, X., et al. Role of homocysteine in the development and progression of Parkinson’s disease. Annals of Clinical and Translational Neurology 7, 2332–2338 (2020).

34. Huang, Y.-S., Ogbechi, J., Clanchy, F.I., Williams, R.O. & Stone, T.W. IDO and Kynurenine Metabolites in Peripheral and CNS Disorders. Frontiers in Immunology 11(2020).

35. Darcy, C.J., et al. An observational cohort study of the kynurenine to tryptophan ratio in sepsis: association with impaired immune and microvascular function. PLoS One 6, e21185 (2011).

36. Wang, Q., Liu, D., Song, P. & Zou, M.H. Tryptophan-kynurenine pathway is dysregulated in inflammation, and immune activation. Front Biosci (Landmark Ed) 20, 1116–1143 (2015).

37. Davey, A., Elias, M.F., Robbins, M.A., Seliger, S.L. & Dore, G.A. Decline in renal functioning is associated with longitudinal decline in global cognitive functioning, abstract reasoning and verbal memory. Nephrology Dialysis Transplantation 28, 1810–1819 (2013).

38. Haukka, J.K., et al. Metabolomic Profile Predicts Development of Microalbuminuria in Individuals with Type 1 Diabetes. Scientific Reports 8, 13853 (2018).

39. Kraft, M.L. Sphingolipid Organization in the Plasma Membrane and the Mechanisms That Influence It. Front Cell Dev Biol 4, 154 (2016).

40. Nezhadi, A., Sheibani, V., Esmaeilpour, K., Shabani, M. & Esmaeili-Mahani, S. Neurosteroid allopregnanolone attenuates cognitive dysfunctions in 6-OHDA-induced rat model of Parkinson’s disease. Behav Brain Res 305, 258–264 (2016).

41. Peng, F., et al. Low antioxidant status of serum uric acid, bilirubin and albumin in patients with neuromyelitis optica. Eur J Neurol 19, 277–283 (2012).

42. Moccia, M., et al. Increased bilirubin levels in de novo Parkinson’s disease. Eur J Neurol 22, 954–959 (2015).

43. Scigliano, G., et al. Increased plasma bilirubin in Parkinson patients on L-dopa: evidence against the free radical hypothesis? Ital J Neurol Sci 18, 69–72 (1997).

44. Macías-García, D., et al. Increased bilirubin levels in Parkinson’s disease. Parkinsonism Relat Disord 63, 213–216 (2019).

45. Yoo, M.S., et al. Oxidative stress regulated genes in nigral dopaminergic neuronal cells: correlation with the known pathology in Parkinson’s disease. Brain Res Mol Brain Res 110, 76–84 (2003).

46. Schipper, H.M., Song, W., Zukor, H., Hascalovici, J.R. & Zeligman, D. Heme oxygenase-1 and neurodegeneration: expanding frontiers of engagement. J Neurochem 110, 469–485 (2009).

47. Yamamoto, N., et al. Elevation of heme oxygenase-1 by proteasome inhibition affords dopaminergic neuroprotection. J Neurosci Res 88, 1934–1942 (2010).

48. Tuschl, K., et al. Mutations in SLC39A14 disrupt manganese homeostasis and cause childhood-onset parkinsonism-dystonia. Nat Commun 7, 11601 (2016).

49. Williams, B.B., et al. Glycoprotein A33 deficiency: a new mouse model of impaired intestinal epithelial barrier function and inflammatory disease. Dis Model Mech 8, 805–815 (2015).

50. Garcia-Esparcia, P., Hernández-Ortega, K., Ansoleaga, B., Carmona, M. & Ferrer, I. Purine metabolism gene deregulation in Parkinson’s disease. Neuropathol Appl Neurobiol 41, 926–940 (2015).

51. Lorenzl, S., et al. Tissue inhibitors of matrix metalloproteinases are elevated in cerebrospinal fluid of neurodegenerative diseases. J Neurol Sci 207, 71–76 (2003).

52. Hughes, A.J., Daniel, S.E., Ben-Shlomo, Y. & Lees, A.J. The accuracy of diagnosis of parkinsonian syndromes in a specialist movement disorder service. Brain 125, 861–870 (2002).

53. Allocati, N., Masulli, M., Di Ilio, C. & Federici, L. Glutathione transferases: substrates, inihibitors and pro-drugs in cancer and neurodegenerative diseases. Oncogenesis 7, 8 (2018).

54. Shi, M., et al. Identification of glutathione S-transferase pi as a protein involved in Parkinson disease progression. Am J Pathol 175, 54–65 (2009).

55. Alsina, F.C., et al. Lrig1 is a cell-intrinsic modulator of hippocampal dendrite complexity and BDNF signaling. EMBO Rep 17, 601–616 (2016).

56. Hasin, Y., Seldin, M. & Lusis, A. Multi-omics approaches to disease. Genome Biol 18, 83 (2017).

57. Arif, M., et al. iNetModels 2.0: an interactive visualization and database of multi-omics data. Nucleic Acids Res (2021).

58. Birkmayer, J.G., Vrecko, C., Volc, D. & Birkmayer, W. Nicotinamide adenine dinucleotide (NADH)--a new therapeutic approach to Parkinson’s disease. Comparison of oral and parenteral application. Acta Neurol Scand Suppl 146, 32–35 (1993).

59. Kuhn, W., et al. Parenteral application of NADH in Parkinson’s disease: clinical improvement partially due to stimulation of endogenous levodopa biosynthesis. J Neural Transm (Vienna) 103, 1187–1193 (1996).

60. LeWitt, P.A., Li, J., Lu, M., Guo, L. & Auinger, P. Metabolomic biomarkers as strong correlates of Parkinson disease progression. Neurology 88, 862–869 (2017).

61. Meiser, J., et al. Loss of DJ-1 impairs antioxidant response by altered glutamine and serine metabolism. Neurobiol Dis 89, 112–125 (2016).

62. Lewitt, P.A., et al. 3-hydroxykynurenine and other Parkinson’s disease biomarkers discovered by metabolomic analysis. Mov Disord 28, 1653–1660 (2013).

63. Zhao, H., et al. Potential biomarkers of Parkinson’s disease revealed by plasma metabolic profiling. J Chromatogr B Analyt Technol Biomed Life Sci 1081-1082, 101–108 (2018).

64. Covarrubias, A.J., Perrone, R., Grozio, A. & Verdin, E. NAD(+) metabolism and its roles in cellular processes during ageing. Nat Rev Mol Cell Biol 22, 119–141 (2021).

65. Gelfin, E., et al. D-serine adjuvant treatment alleviates behavioural and motor symptoms in Parkinson’s disease. Int J Neuropsychopharmacol 15, 543–549 (2012).

66. Shigenaga, M.K., Hagen, T.M. & Ames, B.N. Oxidative damage and mitochondrial decay in aging. Proc Natl Acad Sci U S A 91, 10771–10778 (1994).

67. Ferreira, G.C. & McKenna, M.C. L-Carnitine and Acetyl-L-carnitine Roles and Neuroprotection in Developing Brain. Neurochem Res 42, 1661–1675 (2017).

68. Cherix, A., et al. Metabolic signature in nucleus accumbens for anti-depressant-like effects of acetyl-L-carnitine. Elife 9(2020).

69. Salminen, A., Jouhten, P., Sarajärvi, T., Haapasalo, A. & Hiltunen, M. Hypoxia and GABA shunt activation in the pathogenesis of Alzheimer’s disease. Neurochem Int 92, 13–24 (2016).

70. Supandi, F. & van Beek, J. Computational prediction of changes in brain metabolic fluxes during Parkinson’s disease from mRNA expression. PLoS One 13, e0203687 (2018).

71. Chew, H., Solomon, V.A. & Fonteh, A.N. Involvement of Lipids in Alzheimer’s Disease Pathology and Potential Therapies. Frontiers in physiology 11, 598–598 (2020).

72. Castellanos, D.B., Martín-Jiménez, C.A., Rojas-Rodríguez, F., Barreto, G.E. & González, J. Brain lipidomics as a rising field in neurodegenerative contexts: Perspectives with Machine Learning approaches. Front Neuroendocrinol 61, 100899 (2021).

73. Yakhine-Diop, S.M.S., et al. Metabolic alterations in plasma from patients with familial and idiopathic Parkinson’s disease. Aging (Albany NY) 12, 16690–16708 (2020).

74. Wyss, M. & Kaddurah-Daouk, R. Creatine and creatinine metabolism. Physiol Rev 80, 1107–1213 (2000).

75. Matthews, R.T., et al. Creatine and cyclocreatine attenuate MPTP neurotoxicity. Exp Neurol 157, 142–149 (1999).

76. Li, Z., et al. The effect of creatine and coenzyme q10 combination therapy on mild cognitive impairment in Parkinson’s disease. Eur Neurol 73, 205–211 (2015).

77. Couzin, J. Testing a Novel Strategy Against Parkinson’s Disease. Science 315, 1778 (2007).

78. Watanabe, A., Kato, N. & Kato, T. Effects of creatine on mental fatigue and cerebral hemoglobin oxygenation. Neurosci Res 42, 279–285 (2002).

79. Zhong, L.-L., Song, Y.-Q., Tian, X.-Y., Cao, H. & Ju, K.-J. Level of uric acid and uric acid/creatinine ratios in correlation with stage of Parkinson disease. Medicine (Baltimore) 97, e10967–e10967 (2018).

80. Elias, M.F., Dore, G.A. & Davey, A. Kidney disease and cognitive function. Contrib Nephrol 179, 42–57 (2013).

81. Hare, T.A., Vanna, S., Beasley, B., Chambers, R. & Vogel, W.H. Amino acid and dopa levels in plasma and urine from L-dopa-treated patients with Parkinson’s disease. J Lab Clin Med 77, 319–325 (1971).

82. Manyam, B.V., Ferraro, T.N. & Hare, T.A. Cerebrospinal fluid amino compounds in Parkinson’s disease. Alterations due to carbidopa/levodopa. Arch Neurol 45, 48–50 (1988).

83. Hatano, T., Saiki, S., Okuzumi, A., Mohney, R.P. & Hattori, N. Identification of novel biomarkers for Parkinson’s disease by metabolomic technologies. J Neurol Neurosurg Psychiatry 87, 295–301 (2016).

84. Wang, H., et al. High urea induces depression and LTP impairment through mTOR signalling suppression caused by carbamylation. EBioMedicine 48, 478–490 (2019).

85. Wikoff, W.R., et al. Metabolomics analysis reveals large effects of gut microflora on mammalian blood metabolites. Proceedings of the National Academy of Sciences 106, 3698 (2009).

86. Mangalam, A., et al. Profile of Circulatory Metabolites in a Relapsing-remitting Animal Model of Multiple Sclerosis using Global Metabolomics. J Clin Cell Immunol 4(2013).

87. Grant, S.M. & DeMorrow, S. Bile Acid Signaling in Neurodegenerative and Neurological Disorders. Int J Mol Sci 21(2020).

88. Graham, S.F., et al. Biochemical Profiling of the Brain and Blood Metabolome in a Mouse Model of Prodromal Parkinson’s Disease Reveals Distinct Metabolic Profiles. J Proteome Res 17, 2460–2469 (2018).

89. Burstein, S.H. N-Acyl Amino Acids (Elmiric Acids): Endogenous Signaling Molecules with Therapeutic Potential. Mol Pharmacol 93, 228–238 (2018).

90. Palacios, N., et al. Circulating Plasma Metabolites and Cognitive Function in a Puerto Rican Cohort. Journal of Alzheimer’s disease : JAD 76, 1267–1280 (2020).

91. Błaszczyk, J.W. The Emerging Role of Energy Metabolism and Neuroprotective Strategies in Parkinson’s Disease. Front Aging Neurosci 10, 301–301 (2018).

92. Brandão, P.R.P., et al. Cognitive impairment in Parkinson’s disease: A clinical and pathophysiological overview. J Neurol Sci 419, 117177 (2020).

93. Poston, K.L. & Eidelberg, D. FDG PET in the Evaluation of Parkinson’s Disease. PET Clin 5, 55–64 (2010).

94. Zhang, N., et al. Comprehensive serum metabolic and proteomic characterization on cognitive dysfunction in Parkinson’s disease. Ann Transl Med 9, 559 (2021).

95. Emre, M., et al. Rivastigmine for dementia associated with Parkinson’s disease. N Engl J Med 351, 2509–2518 (2004).

96. Solari, N., Bonito-Oliva, A., Fisone, G. & Brambilla, R. Understanding cognitive deficits in Parkinson’s disease: lessons from preclinical animal models. Learn Mem 20, 592–600 (2013).

97. Antonini, A. & Cilia, R. Behavioural adverse effects of dopaminergic treatments in Parkinson’s disease: incidence, neurobiological basis, management and prevention. Drug Saf 32, 475–488 (2009).

