## supplementary appendix for "Combined Metabolic Activators Improve Cognitive Functions without Altering Motor Scores in Parkinson’s Disease"

### TRIAL DESIGN

### OBJECTIVES AND RATIONALE

The **primary objective** was to assess the clinical difference in motor and cognitive functions between the subjects receiving twelve-weeks treatment either with metabolic cofactors supplementation or placebo.

The primary analyses were on the difference in Unified Parkinson's Disease Rating Scale (UPDRS) scores and The Montreal Cognitive Assessment (MoCA) test scores between the placebo and the treatment arms in PD patients.

The **secondary objectives** were:

(1) to determine the behavioural symptoms of PD patients were evaluated via Neuropsychiatric Inventory NPI.

(2) to assess the tolerability and safety profile of co-factor supplementation;

(3) to examine the additional efficacy parameters including metabolomic and proteomic analysis.

##### COMPOSITION AND DOSAGES

| **Metabolites** | **Molar Weight** | **Molar Ratio** | **Gram in cocktail** | **Molar** | **Ratio** |
| --- | --- | --- | --- | --- | --- |
| L-carnitine (C7H15NO3)  Molar mass 161.199 g/mole | 161.20 | 4 | 5.1 | 0.031 | 1 |
| N-acetyl-L-cysteine (NAC) (C5H9NO3S)  Molar mass 163.195 g/mole | 163.20 | 4 | 5.1 | 0.031 | 0.99 |
| Nicotinamide riboside (NR) (C11H15N2O5+)  Molar mass 255.25 g/mole | 255.25 | 1 | 2 | 0.0078 | 0.25 |
| Serine (C3H7NO3)  Molar mass 105.09 g/mole | 105.09 | 30 | 24.7 | 0.234 | 7.67 |

This double-blind, randomized, placebo-controlled, trial aimed to establish metabolic improvements in PD subjects by dietary supplementation with cofactors N-acetylcysteine, L-carnitine tartrate, nicotinamide riboside and serine. The subjects took a mixture of cofactors or matching placebo as powder dissolved in water by mouth.

The investigational drug products, the mixture of the four co-factors and the placebo was produced in Turkey according to GMP regulations by Pharmactive Company ([www.pharmactive.com.tr/en/anasayfa.html](http://www.pharmactive.com.tr/en/anasayfa.html)) according to EU standards and packed according to GMP rules with the appropriate dosage. Investigational product and placebo were produced as soluble powders and were packed in 60 mL HDPE plastic bottles with screw caps. Bottles were packed in carton boxes with appropriate inner and outer labelling according to applicable local regulations. Pharmactive has European GMP certificate. Specifications of each supplier of the individual GMP grade co-factors included in the drug product were enclosed. The investigational test product and placebo were administered in white plastic bottles (containing dry powder) with a desiccant cap.

- Stability tests performed by ChromaDex in USA show that the cocktail of cofactors is stable at 25°C for at least three months in the plastic bottles.
- The cofactors were administered as a powder with strawberry aroma to be dissolved in 200 ml of preferably cold still drinking water and be consumed within 5 minutes.
- As placebo, sorbitol^[1]^ (5 g) flavoured with strawberry aroma and colouring agent were given.

#### STUDY POPULATION

The study population were consisting of 48 Parkinson’s patients. Eligible subjects signed an informed consent, meet all inclusion criteria and had none of the exclusion criteria listed below.

##### **Inclusion criteria**

I-1. Men and women diagnosed with Parkinson’s Disease (Hoehn Yahr 2-4, age >40 years).
I-2. Patients with stable treatments and clinical course

##### **Exclusion criteria**

E-1. Inability or unwillingness to give written informed consent
E-2. History of stroke, severe brain trauma, toxic drug exposure
E-3. Neurological examination which indicate to Parkinson-Plus syndrome (i.e., pyramidal, cerebellar and autonomic dysfunction findings and gaze paralysis) for PD
E-4. Uncontrolled Type 1 or type 2 diabetes
E-5. Diarrhea (defined as more than 2 stool per day) within 7 days before enrolment
E-6. Chronic kidney disease with an estimated glomerular filtration rate <60 ml/min/1.73m2
E-7. Significant cardiovascular co-morbidity (i.e. heart failure)
E-8. Patients with active bronchial asthma
E-9. Patients with phenylketonuria (contraindicated for NAC)
E-10. Patients with histamine intolerance
E-11. Clinically significant TSH level outside the normal range (0.04-6 mU/L)
E-12. Known allergy for substances used in the study
E-13. Concomitant medication use:
      - Self-administration of dietary supplements such as any vitamins, omega-3 products, or plant stanol/sterol products within 1 week
      - Use of an antimicrobial agent in the 4 weeks preceding randomization
E-14. Active smokers consuming >10 cigarettes/day
E-15. Alcohol consumption over 192 grams for men and 128 grams for women per week
E-16. Patients considered as inappropriate for this study for any reason
E-17. Active participation in another clinical study
E-18. Women with potential to give birth. Post-menopausal women, women who had hysterectomy or women with acceptable medical proof shoving that she can’t get pregnant according to the investigator’s decision may be enrolled to the study.

#### RANDOMIZATION AND BLINDING

The study subjects were randomized on a 2:1 basis to the cofactor mixture or placebo. A web-based randomization system was used to assign a randomization code for each patient. Investigator or other responsible person at the investigational site were able to enter the web-based randomization system specific to the study through assigned username and password. After entering patient-related information (patient number, date of birth, patient initials), the system was provided randomization code for the future use by investigators. This randomization code was entered into the electronic case report form (e-CRF).

#### DOSAGE AND ADMINISTRATION

Subjects in active treatment were receive dietary supplementation with N-acetylcysteine, L-carnitine tartrate, nicotinamide riboside, and serine, administered as a mixture. Half dosage of the co-factors was given for two weeks (one dose taken just after dinner), and full dosage for 8 weeks (two equal doses taken just after breakfast and dinner).

The dosage of the supplements was as follows:

|  | **Weeks 1-4 (28 days)** | **Weeks 5-12 (56 days)** |
| --- | --- | --- |
| L-Carnitine tartrate | 3.73 g/day | 7.46 g/day |
| N-Acetylcysteine | 2.55/day | 5.1 g/day |
| Nicotinamide riboside | 1 g/day | 2 g/day |
| Serine | 12.35 g/day | 24.7 g/day |

Patients who cannot tolerate taking full dose may continue the study with half dose (i.e. one dose taken just after dinner). Patients who cannot tolerate the study agents with half dose were withdrawn from the study.

##### SCREENING VISIT

**Screening visit (Days -7 to -1).** The subjects got the informed consent form and oral information on the study protocol, including omics studies, number of visits and procedures to be performed in each visit. After the subject has given written informed consent to participate in the study, the following data were obtained in the screening visit:

- Demographic data
- Medical history including associated conditions, previous operations, smoking and alcohol consumption and concomitant drug usage
- Physical examination including body weight, height, waist circumference, hip circumference, blood pressure and pulse rate
- Neurological examination to stage PD patients
- Functional Medicine Assessment
- Review of inclusion/exclusion criteria
- Laboratory safety parameters including complete blood count (blood cells, haemoglobin), alkaline phosphatase (ALP), alanine aminotransferase (ALT), Aspartate aminotransferase (AST), total cholesterol, high density lipoprotein-cholesterol (HDL-C), low density lipoprotein-cholesterol (LDL-C), triglycerides, creatinine, urea.
- Advanced serum metabolomics with generation of untargeted metabolomics data.
- The motor and cognitive functions of PD patients will be evaluated via UPDRS and MoCA, respectively.
- The behavioural functions of PD patients were evaluated via NPI.
- Information on prior and concomitant medications were collected
- Adverse events were questioned.

##### VISIT 1

**Randomization visit (Day 0).** Eligible study subjects were randomized to active therapy or placebo groups and study agents were dispensed. Additionally, surgical/medical history as well as information on prior and concomitant medication were collected and physical/neurological examination were repeated.

They were asked to take the first dose at the hospital and be observed for development of adverse events. The changes in concomitant medications were evaluated. Patients who can tolerate the study agents were start to take half dose of co-factors supplementation (i.e., one administration daily just after dinner) for two weeks. Treatment compliance and adverse event questioning were performed by the study nurse by weekly telephone contacts.

##### VISIT 2

**On-treatment first visit (Day 28).** The subjects were come to the study center for complete evaluation including and advers events recording. Clinical and physical examination, determination of the motor, cognitive and behavioural functions using clinical scales, laboratory safety parameters, proteomic and metabolomic were repeated as in Visit 1.

Clinical scales included UPDRS and MoCA. NPI were applied in order to evaluate behavioural symptoms.

After the visit 3, subjects were start to take full dose of final concentrations (*i.e.,* 2 dosages daily taken just after breakfast and after dinner). Treatment compliance and adverse event questioning were performed by the study nurse by weekly telephone contacts.

VISIT 3

**End of study, on-treatment second visit (Day 84).** The subjects were return to the study center after eight weeks to evaluate efficacy, tolerability and safety.

At the end of the treatment, all procedures including clinical and physical examination, adverse events recording, determination of the motor, cognitive and behavioral functions using clinical scales, biochemical, metabolomic and proteomic analysis were repeated as in Visit 1. Treatment compliance and development of any adverse events were questioned during this visit.

|  | **Pre-treatment Phase** | **Treatment Period** | | |
| --- | --- | --- | --- | --- |
| **Visits** | **Screening** | **Visit 1** | **Visit 2** | **Visit 3** |
| **Days** | **Days -7 to -1** | **Day 0** | **Day 28** | **Day 84** |
| Informed consent | X |  |  |  |
| Demographic data | X |  |  |  |
| Surgical & medical history | X | X | X | X |
| Physical/neurological examination^1^ | X | X | X | X |
| Inclusion/Exclusion criteria | X | X |  |  |
| Functional Med. Evaluation | X |  |  | X |
| **Laboratory tests** |  |  |  |  |
| Blood sample collection^2^ | X |  | X | X |
| **Efficacy and safety evaluation** |  |  |  |  |
| Laboratory safety parameters ^3^ | X |  | X | X |
| Serum metabolomics & lipid analysis ^4^ | X |  | X | X |
| **Clinical and Radiological assessments** |  |  |  |  |
| The Montreal Cognitive Assessment (MoCA) ^5^ | X |  | X | X |
| Unified Parkinson's Disease Rating Scale (UPDRS) ^6^ | X |  | X | X |
| Neuropsychiatric Inventory (NPI) ^7^ | X |  | X | X |
| **Study drug administration** |  |  |  |  |
| Randomization ^8^ |  | X |  |  |
| Half dosage ^9^ |  | X ^10^ |  |  |
| Full dosage ^11^ |  |  | X |  |
| Monitoring of compliance ^12^ |  | X | X | X |
| Prior & concomitant medication | X | X | X | X |
| AE and SAE ^13^ | X | X | X | X |

1. Physical examination included body weight, height, body mass index, vital signs, waist and hip circumferences measurements.

2. At visits 1, 2 and 3, whole blood samples were collected for omics analysis.

3. Laboratory safety parameters included; complete blood count (blood cells, hemoglobin), alkaline phosphatase (ALP), alanine aminotransferase (ALT), aspartate aminotransferase (AST), total cholesterol, high density lipoprotein-cholesterol (HDL-C), low density lipoprotein-cholesterol (LDL-C), triglycerides, creatinine, urea

4. Serum metabolic and proteomic analysis included generation of untargeted metabolomics and proteomics data.

5. Cognitive evaluation scale for PD patients

6. Motor evaluation scale for PD patients

7. Behavioural evaluation scale for PD patients

8. Eligible study subjects at screening were offered to be enrolled in the study and invited back to the clinic within the following week for randomization and to take the first dose at the hospital.

9. One dose taken just after dinner

10. The study participants were observed for the development of any allergic reactions or intolerance after taking the first dose at the hospital.

11. After Visit 2, subjects were switch to full dose and start to receive two equal doses just after breakfast and dinner

12. Compliance and adverse events were assessed by weekly phone contact with the study nurse.

13. Adverse events (AE) and serious adverse events (SEA) were monitored continuously and all AEs that occur at any time during the study were reported in Case Report Forms.

#### **SUBJECT COMPLETION/WITHDRAWAL**

A subject was considered as having completed the study if he/she has completed all assessments at the End of Treatment Visit and has been followed up until 12 weeks after initiation of the study drugs.

Subjects were informed that they have the right to discontinue from the study at any time, without prejudice to their medical care.

Any patient may be withdrawn from the study for reasons beneficial to his/her welfare and/or upon his/her request. A subject should be discontinued from the study if

- the investigator decides that for safety reasons (i.e., clinically significant adverse event and/or laboratory abnormalities) it is in the best interest of the subject to stop treatment
- the subject becomes pregnant
- the subject starts taking concomitant medications listed among the exclusion criteria (e.g. other dietary supplements, etc.)

Major protocol violations, non-compliance and administrative issues may be reasons for early discontinuation of the study agents.

The reason(s) for a subject’s discontinuation were recorded on the CRF.

#### ASSESMENTS AND OUTCOME PARAMETERS

**Assessment of the efficacy**

The primary endpoint was the difference in cognitive and motor functions between subjects taking co-factor supplementation and placebo after 12 weeks of treatment in PD patients. In this respect, PD patients were received UPDRS, MoCA and NPI in order to evaluate the functions of motor, cognition and behavior respectively.

**Other assessments**

1. *Laboratory safety tests*

Laboratory safety parameters including complete blood count (blood cells, haemoglobin), alkaline phosphatase (ALP), alanine aminotransferase (ALT), Aspartate aminotransferase (AST), gamma-glutamyl transferase (GGT), bilirubin, albumin, creatinine kinase (CK), total cholesterol, high density lipoprotein-cholesterol (HDL-C), low density lipoprotein-cholesterol (LDL-C), triglycerides, creatinine, urea, urate, glucose, sodium (Na), potassium (K), insulin, HbA1c and thyroid-stimulating hormone (TSH) were assessed in the local laboratory of the Istanbul Medipol and Alanya Alaaddin Keykubat University Hospitals. 5 mL of blood were collected for this purpose.

2. *Plasma omics analysis*

Blood were centrifugated, aliquoted and stored at -80◦C. After storage, for analysis all plasma samples were transferred on dry ice to Sweden (preferentially by WorldCourier) for further analyses. Biomarkers were analysed by proximity extension and proximity ligation technologies (PEA and PLA) providing assays with high specificity and sensitivity in complex biological matrices. Metabolomics (untargeted) and proteomics (untargeted) were analysed by mass spectrometry technology. 5 mL of blood were collected for this purpose. Whole genome sequencing was not performed.

**Assessment of safety**

We assessed the patient compliance and carefully follow if there is any side effect of the dietary supplementation. Vital signs (blood pressure, pulse), weight and laboratory safety parameters (detailed above) were followed at each visit. Any symptoms of intestinal discomfort or other side effects were carefully recorded. The subjects were informed to contact (by phone or text message) the investigators immediately if they experience any symptoms of discomfort or any side effects during the intervention period.

### REPORTING ADVERSE EVENTS AND SAFETY ANALYSIS

**Definitions**

An adverse event is any noxious and unintended change in the patient’s profile involving function, structure, or chemistry that occurs during the study, regardless of diet relationship, including any undercurrent illness, injury, toxicity, sensitivity, or sudden death.

A serious adverse event is defined as any experience that is fatal, immediately life threatening, severely or permanently disabling, results in a congenital anomaly, requiring or prolonging hospitalization, or is an important medical event that may jeopardize the patient and may require medical or surgical intervention to prevent one of the other outcomes.

An unlisted adverse event is any event having the nature or severity not consistent with the applicable product reference safety information.

An adverse event is considered associated with the use of the drug if the attribution is possible, probable, or very likely according to the definitions listed below:

- Not related - An adverse event that is not related to the use of the drug.
- Doubtful - An adverse event for which an alternative explanation is more likely, e.g., concomitant drug(s), concomitant disease(s), or the relationship in time suggests that a causal relationship is unlikely.
- Possible - An adverse event that might be due to the use of the drug. An alternative explanation, e.g., concomitant drug(s), concomitant disease(s), is inconclusive. The relationship in time is reasonable; therefore, the causal relationship cannot be excluded.
- Probable - An adverse event that might be due to the use of the drug. The relationship in time is suggestive (e.g., confirmed by dechallenge). An alternative explanation is less likely, e.g., concomitant drug(s), concomitant disease(s).
- Very likely - An adverse event that is listed as a possible adverse reaction and cannot be reasonably explained by an alternative explanation, e.g., concomitant drug(s), concomitant disease(s). The relationship in time is very suggestive (e.g., it is confirmed by dechallenge and rechallenge).

**Procedures**

The intent-to-treat analysis were used to evaluate issues of safety. Safety evaluations were based on adverse events, scheduled physical examinations, vital signs (blood pressure, heart rate) and clinical laboratory tests.

All adverse events that occur at any time during the study were reported in the patient’s CRF. All adverse events, regardless of seriousness, severity, or presumed relationship to study therapy, were recorded using medical terminology in the source document and in the CRF. Investigators were also record their opinion concerning the relationship of the adverse event to study agent.

Any serious adverse event was reported to the sponsor within 24 hours of their knowledge of the event. All initial reports of pregnancy were reported to the sponsor by the investigators within 24 hours after of they were informed on the event.

All adverse events were followed until resolution or improvement of the event. All adverse events that have not resolved by the end of the study, or that have not resolved upon discontinuation of the study agent were followed until any of the following occurs:

–	the event resolves

–	the event stabilizes

–	the event returns to baseline, if a baseline value is available

–	the event can be attributed to agents other than the study drug or to factors unrelated to study conduct

–	when it becomes unlikely that any additional information can be obtained (subject or health care practitioner refusal to provide additional information, lost to follow-up after demonstration of due diligence with follow-up efforts)

The investigators and the sponsor assume responsibility for appropriate reporting of adverse events to the Competent Authorities. The investigators must report these events to the Institutional Review Board of Istanbul Medipol University.

### ETHICAL ISSUES

- Participation in the study is entirely voluntary for the research subjects.
- All procedures and possible hazards, risks and discomforts were explained to the subjects both orally and in writing. We have years of experience in intensive protocols like this, with great success.
- All study materials including patient information and the laboratory results were kept confidential.
- Study participants were refunded for the discomfort, and also travel costs to the research sites were covered from research funds.
- In case of a patient accident, the patients are under the normal patient insurance and can have refunds through hospital’s normal policy.
- Data of study subjects who have dropped out from the study were remain in the data file unless study subjects request their data to be excluded.
- During the study the investigators (or sponsor where required) were send the following documents to the IEC/IRB for their review and approval, where appropriate:
  - protocol amendments
  - revision(s) to informed consent form and any other written materials to be provided to subjects
  - Investigator’s Brochure amendments or new edition(s)
  - reports of adverse events that are serious, unlisted, and associated with the investigational drug
  - new information that may adversely affect the safety of the subjects or the conduct of the study
  - deviations from or changes to the protocol to eliminate immediate hazards to the subjects
  - report of deaths of subjects under the investigator's care
  - notification if a new investigator is responsible for the study at the site

**Regulatory Approval/Notification**

This protocol and any amendment(s) were submitted to the Ministry of Health of Turkey. The study initiated when regulatory requirements were met.

**Monitoring**

The sponsor performed on-site monitoring visits as frequently as necessary. At all visits, the monitor compared the data entered into the CRFs with the hospital or clinic records (source documents). The nature and location of all source documents were identified to ensure that all sources of original data required to complete the CRF are known to the sponsor and investigational staff and are accessible for verification by the sponsor site contact.

Direct access to source documentation (medical records) must be allowed for the purpose of verifying that the data recorded in the CRF are consistent with the original source data. The monitor met with the investigator on a regular basis during the study to provide feedback on the study conduct.

1. Sorbitol is widely used due to its solubility in water. It’s approved by the U.S. Food and Drug Administration (FDA). See link: [Inactive Ingredients in FDA Approved Drugs](http://www.accessdata.fda.gov/scripts/cder/iig/index.cfm).
